# Genetic predisposition to psychiatric disorders and risk of COVID-19

**DOI:** 10.1101/2021.02.23.21251866

**Authors:** Wenwen Chen, Yu Zeng, Chen Suo, Huazhen Yang, Yilong Chen, Can Hou, Yao Hu, Zhiye Ying, Yajing Sun, Yuanyuan Qu, Donghao Lu, Fang Fang, Unnur A. Valdimarsdóttir, Huan Song

## Abstract

**Background:** Pre-pandemic psychiatric disorders have been associated with an increased risk of COVID-19. However, the underlying mechanisms remain unknown, e.g. to what extent genetic predisposition to psychiatric disorders contributes to the observed association.

**Methods:** The analytic sample consisted of white British participants of UK Biobank registered in England, with available genetic data, and alive on Jan 31, 2020 (i.e., the start of the COVID-19 outbreak in the UK) (n=346,554). We assessed individuals’ genetic predisposition to different psychiatric disorders, including substance misuse, depression, anxiety, and psychotic disorder, using polygenic risk score (PRS). Diagnoses of psychiatric disorders were identified through the UK Biobank hospital inpatient data. We performed a GWAS analysis for each psychiatric disorder in a randomly selected half of the study population who were free of COVID-19 (i.e., the base dataset). For the other half (i.e., the target dataset), PRS was calculated for each psychiatric disorder using the discovered genetic variants from the base dataset. We then examined the association between PRS of each psychiatric disorder and risk of COVID-19, or severe COVID-19 (i.e., hospitalization and death), using logistic regression models. The ascertainment of COVID-19 was through the Public Health England dataset, the UK Biobank hospital inpatient data and death registers, updated until July 26, 2020. For validation, we repeated the PRS analyses based on publicly available GWAS summary statistics.

**Results:** 155,988 participants (including 1,451 COVID-19 cases), with a mean age of 68.50 years at COVID-19 outbreak, were included for PRS analysis. Higher genetic liability forwards psychiatric disorders was associated with increased risk of both any COVID-19 and severe COVID-19, especially genetic risk for substance misuse and depression. The adjusted odds ratios (ORs) for any COVID-19 were 1.15 (95% confidence interval [CI] 1.02-1.31) and 1.26 (1.11-1.42) among individuals with a high genetic risk (above the upper tertile of PRS) for substance misuse and depression, respectively, compared with individuals with a low genetic risk (below the lower tertile). Largely similar ORs were noted for severe COVID-19 and similar albeit slightly lower estimates using PRSs generated from GWAS summary statistics from independent samples.

**Conclusion:** In the UK Biobank, genetic predisposition to psychiatric disorders was associated with an increased risk of COVID-19, including severe course of the disease. These findings suggest the potential role of genetic factors in the observed phenotypic association between psychiatric disorders and COVID-19, underscoring the need of increased medical surveillance of for this vulnerable population during the pandemic.

## Introduction

With over 110 million infected people and 2.4 million related deaths, the coronavirus disease 2019 (COVID-19) pandemic, caused by severe acute respiratory syndrome coronavirus 2 (SARS-CoV-2) virus, has led to an unprecedented crisis worldwide^1^. Evidence suggest that individuals have varying propensity for being infected with COVID-19 and that infected patients demonstrate heterogeneous outcomes^2^, identification of populations with increased susceptibility to the disease, especially ones prone to severe disease course, is critical for optimizing preventive measures.

Previous studies have reported an increased risk of infections, including life-threatening infections, among individuals with psychiatric disorders^3,4^. Likewise, after the COVID-19 outbreak, accumulating evidence revealed that psychiatric disorders^5^, such as depression^2,6^, schizophrenia^7^, and substance abuse^8^ were also associated with an elevated risk of COVID-19, possibly through similar mechanisms as those leading to other infections^9^. Beside the immune dysfunction as widely observed among individuals with psychiatric illness^10^, other explanations might include unfavorable lifestyle, such as smoking and physical inactivity^11^. Furthermore, one recent investigation suggested a shared genetic vulnerability to both psychiatric disorders and infection, reporting a strong genetic association between having at least one psychiatric diagnosis and the occurrence of infection^12^. However, to our knowledge, no study has so far explored whether the genetic predisposition to psychiatric disorders contributes to susceptibility for COVID-19 infection and severe disease course.

Based on the results of genome-wide association studies (GWAS), a polygenic risk score (PRS), or the sum of all risk alleles weighted by the effect size of each variant, can be generated and represent an individual’s overall genetic risk for a given disease such as psychiatric disorders. It can further be used to predict risk of developing a second disease outcome, and thereby illustrate the genetic association between a disease pair^13–15^.

As a continuation of our previous study that demonstrated a robust association between pre-pandemic psychiatric disorders and COVID-19 risk^9^, we here aimed to explore potential underlying mechanisms by testing whether genetic predisposition to psychiatric disorders is associated with risk of SARS-CoV-2 infection and progressive COVID-19 illness using the UK Biobank databases.

## Methods

### Study design

#### The UK Biobank

Our study is based on data from the large-scale prospective cohort of UK Biobank, which enrolled 502,507 individuals aged between 40 and 69 years across the UK during 2006-2010. The genotyping data were obtained from 488,377 blood samples collected at baseline for each participant. They were assayed using the Applied Biosystems UK BiLEVE and UK Biobank Axiom Array^16^. After the quality control following the UK Biobank pipeline, genotype imputation was further completed using the Haplotype Reference Consortium (HRC) and UK10K haplotype resource as reference panels^16^. Kinship coefficient and principal components (PCs), calculated using the KING tool, were also provided by the UK Biobank. Details about the UK Biobank quality control pipeline and imputation methods have been described previously^16^. The final quality controlled and imputed genotypes dataset was the basis of the present analysis, containing more than 93 million autosomal SNPs for 346,554 individuals.

Phenotypic data such as sex and birth year were collected at recruitment using questionnaire. Health-related outcomes were obtained through periodically linked data from multiple national datasets, including death registries and inpatient hospital data from across England, Scotland, and Wales^16^. After the global outbreak of COVID-19, the UK Biobank has also been linked to Public Health England (PHE), where results of COVID-19 tests by RT-PCR (RdRp gene assay) from oral swabs were documented since March 16, 2020^17^.

The UK Biobank collected all data after written informed consent obtained from each participant and the study has full ethical approval from the NHS National Research Ethics Service (16/NW/0274). This present study was also approved by the biomedical research ethics committee of West China Hospital (2020.661).

#### Ascertainment of psychiatric disorders and COVID-19

To keep consistent with our previous analysis of phenotypic association^9^, we used the same approach for the ascertainment of psychiatric disorders and COVID-19 in the present study. Briefly, we defined five broad diagnostic categories of psychiatric disorders, including substance misuse, depressive disorders, anxiety disorders, psychotic disorders, and stress-related disorders, based on hospital admissions with a diagnosis of these disorders according to International Classification of Diseases, Tenth Revision (ICD-10) or ICD-9 codes in the UK Biobank inpatient hospital data (Supplementary Table1) before Jan 31, 2020. The identification of COVID-19 was according to any positive result from the PHE dataset, the diagnosis in the UK Biobank inpatient hospital data, or a cause of death based on the UK Biobank mortality data (ICD codes shown in Supplementary Table1), updated until July 26, 2020. COVID-19 cases from the latter two resources, i.e., hospitalization or death record, were considered as ‘sever cases’.

#### PRS for psychiatric disorders

The genetic analysis was conducted among white British UK Biobank participants who were registered in England at recruitment, alive and trackable on Jan 31, 2020, and having available genetic data (n=346,554). Standard GWAS quality control was performed. Briefly, we restricted our analysis to the autosomal biallelic SNPs and removed variants with a call rate <98%, a minor allele frequency <0.01, or deviation from Hardy–Weinberg equilibrium (P < 10^−6^). We then removed individuals having genotyping rate <98% and outlier samples based on abnormal heterozygosity level, leaving 340,632 participants for further analysis. Details of the quality control are summarized in Supplementary Figure1.

Due to the human heterogeneity and thereby possibly limited portability of PRS between populations, even within those with similar ancestries^18^, we performed a GWAS followed by the PRS analysis for each type of the studied psychiatric disorders by splitting the UK Biobank data into a base and target dataset (study design in Figure 1). To avoid the influence of the phenotypic association (e.g., between depression and COVID-19) on the identification of genetic background for the exposure trait (e.g., depression), in the first step, we removed all individuals with confirmed COVID-19 (n=1,451). Then performed GWAS for each trait in a subsample of the study population, namely 50% of the participants randomly selected from the study population (i.e., the base dataset). Second, we calculated a PRS for each exposure trait for the remaining participants (i.e., the target dataset)^19^, as the weighted sum of the risk alleles based on the summary statistics derived from the GWAS results of the base dataset. Here, the summary statistics referred to the effect sizes and standard errors for the variants. We computed the PRS under ten p value thresholds (i.e., 5 × 10^−8^, 1 × 10^−6^, 1 × 10^−4^, 1 × 10^−3^, 0.05, 0.1, 0.2, 0.3, 0.4 and 0.5).

**Figure 1.**
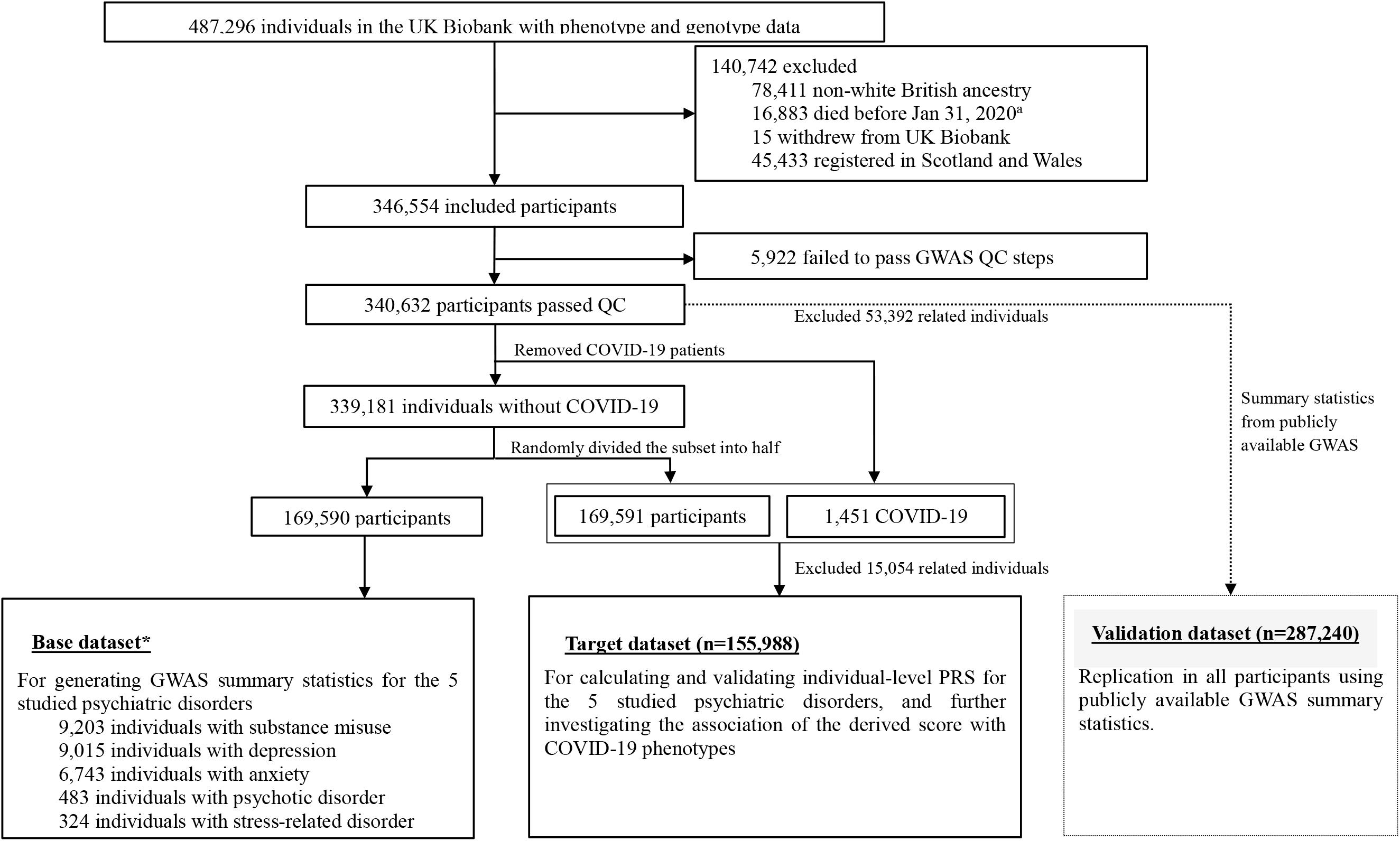
Study design. ^a^ The first COVID-19 case was diagnosed on Jan 31, 2020 in the UK *Removed related individuals prior to each GWAS analysis, with the principle of prioritizing the stay of individuals with the corresponding phenotype

Related individuals, up to the third degree (i.e., kinship coefficients >0.044)^20^, were removed prior to each GWAS or PRS analysis, with the principle of prioritizing the stay of individuals with the corresponding phenotype, if any. Furthermore, in a validation dataset including all eligible and unrelated participants (n=287,240), we also generated PRS for the above psychiatric disorders based on summary statistics from publicly available GWAS^21–24^. PLINK (version 1.9) was used for GWAS and PRS calculation.

### Statistical analysis

First, to validate the predictability of PRS on its each psychiatric phenotype, we used the logistic regression model to measure the association between PRS and each psychiatric disorder category in the base dataset, adjusting for sex, birth year, genotyping batch and significant (i.e., p<0.05) PCs for population heterogeneity. Second, in the target dataset, we examined the association between PRS of a specific psychiatric disorder category and the risk of COVID-19, as well as severe COVID-19, using odds ratios (ORs) with 95% CIs derived from logistic regression models, adjusting for the covariates mentioned above. In addition to considering the standardized PRS as a continuous variable, we also divided participants into low, moderate and high genetic risk groups based on the tertile distribution of the PRS and compared the risk of COVID-19 outcomes using the low genetic risk group (i.e., below the lower tertile of PRS) as a reference. The variance explained by PRS was assessed as the difference in variance, as measured by Nagelkerke’s squared (R square) from the full model including the PRS and the basic model adjusting for sex, birth year, genotyping batch and significant PCs. The analysis was repeated for ten PRSs calculated at different thresholds of p value for each psychiatric disorder category, and the one with the highest R square of COVID-19 was reported as the main results.

To test the robustness of our results, we repeated all the analyses in the validation dataset using PRSs based on summary statistics from publicly available GWAS^21–24^. A 2-sided p<0·05 was considered statistically significant. All the analyses were done with R software, version 4.0.

## Results

In total, 340, 632 participants (Figure 1) were included for analyses with the mean age of 68.51 at the time of the COVID-19 outbreak. 154,065 (45.2%) participants were men. Among the 169,590 participants in the base dataset, there were 9,203 (5.43%), 9,015 (5.32%), 6,743 (3.98%), 483 (0.28%), and 324 (0.19%) cases of substance misuse, depression, anxiety, psychotic disorder and stress-related disorders, respectively. The target dataset included 155,988 participants, including 1,451 COVID-19 cases and 1,059 of these cases who were hospitalized or died due to COVID-19 (severe cases).

### PRS for psychiatric disorders

Based on the base dataset, the GWAS results were summarized using Manhattan plot in Supplementary Figure 2. In brief, except for stress-related disorders, the PRSs (as continuous variables) were significantly associated with increased risk of the corresponding psychiatric disorder in the target dataset (Supplementary Tables 2-6). However, the highest variance explained by one standard deviation increase of PRSs was moderate, with ORs ranging between 1.12 (95% CI 1.09-1.15) and 1.17 (95% CI 1.14-1.20).

**Figure 2.**
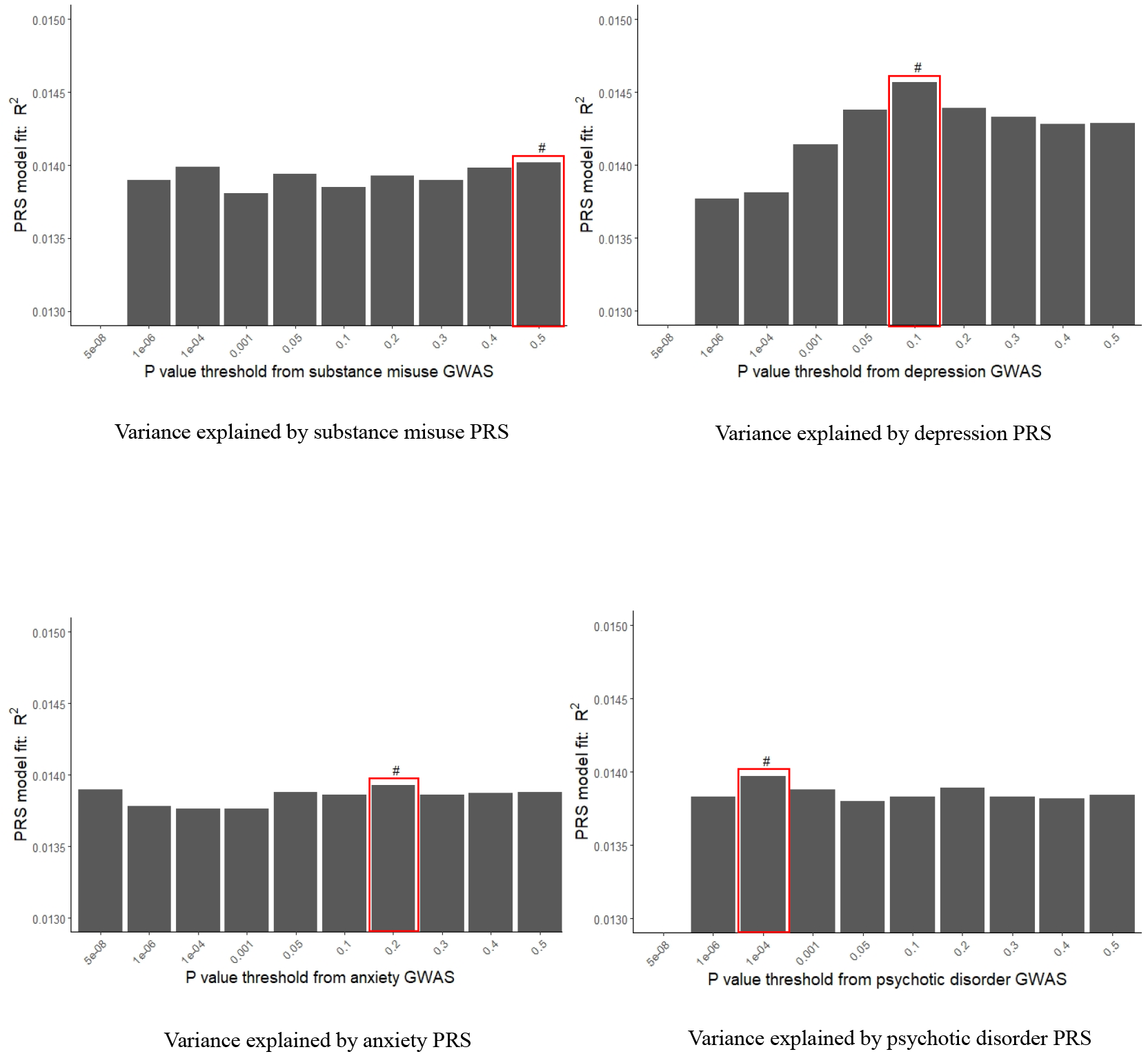
Variance explained for COVID-19 by polygenic risk scores (PRSs) for psychiatric disorder under 10 p value thresholds*. # The p value threshold with the highest variance explained was selected for subsequent analyses. *GWAS summary statistics from UK Biobank base data.

### PRS for psychiatric disorders and COVID-19

Using PRS as a proxy of genetic predisposition to a given psychiatric disorder, we examined the association between genetic risk of a psychiatric disorder and risk of COVID-19, and severe COVID-19, in the target dataset. Given the relatively poor accuracy of the PRS for stress-related disorders, we did not proceed with that PRS in this analysis (Supplementary Table 6). While different p value thresholds were used for PRS calculation, the PRS models with the highest R square, interpreted as the ones with the largest variance explained by the specific psychiatric disorder, were selected as main models for further analyses (Figure 2). Despite the marginally significant association for anxiety, we obtained elevated risks of any COVID-19 in relation to one standard deviation increase in the PRS of all studied psychiatric disorders (Table 1). After adjusting for all covariates, the most pronounced OR was observed for depression, indicating a 10% increased risk of any COVID-19 per standard deviation increase of depression PRS. Analysis of categorized PRS revealed similar results (Figure 3). Notably, for substance misuse and depression, we also observed a dose-response relationship. Compared to individuals with a low genetic risk of depression (PRS<the lower tertile), the adjusted OR was 1.03 (95% CI 0.91-1.18) and 1.26 (95% CI 1.11-1.42) for individuals with moderate (PRS=lower-upper tertile) and high genetic risk (PRS>the upper tertile) of depression, respectively. The corresponding ORs for substance misuse were 1.09 (95% CI 0.96-1.24) and 1.15 (95% CI 1.02-1.31). Regarding severe COVID-19, with lower precision due to smaller case number, we observed stronger associations. For instance, one standard deviation increase in the PRS of depression was associated with 12% (OR, 1.12; 95% CI, 1.06-1.19) increased risk of severe COVID-19 (Table 1). The OR was 1.29 (95% CI 1.12-1.50) among individuals with a high genetic risk of depression, compared with those with a low genetic risk (Figure 3).

**Table 1.**
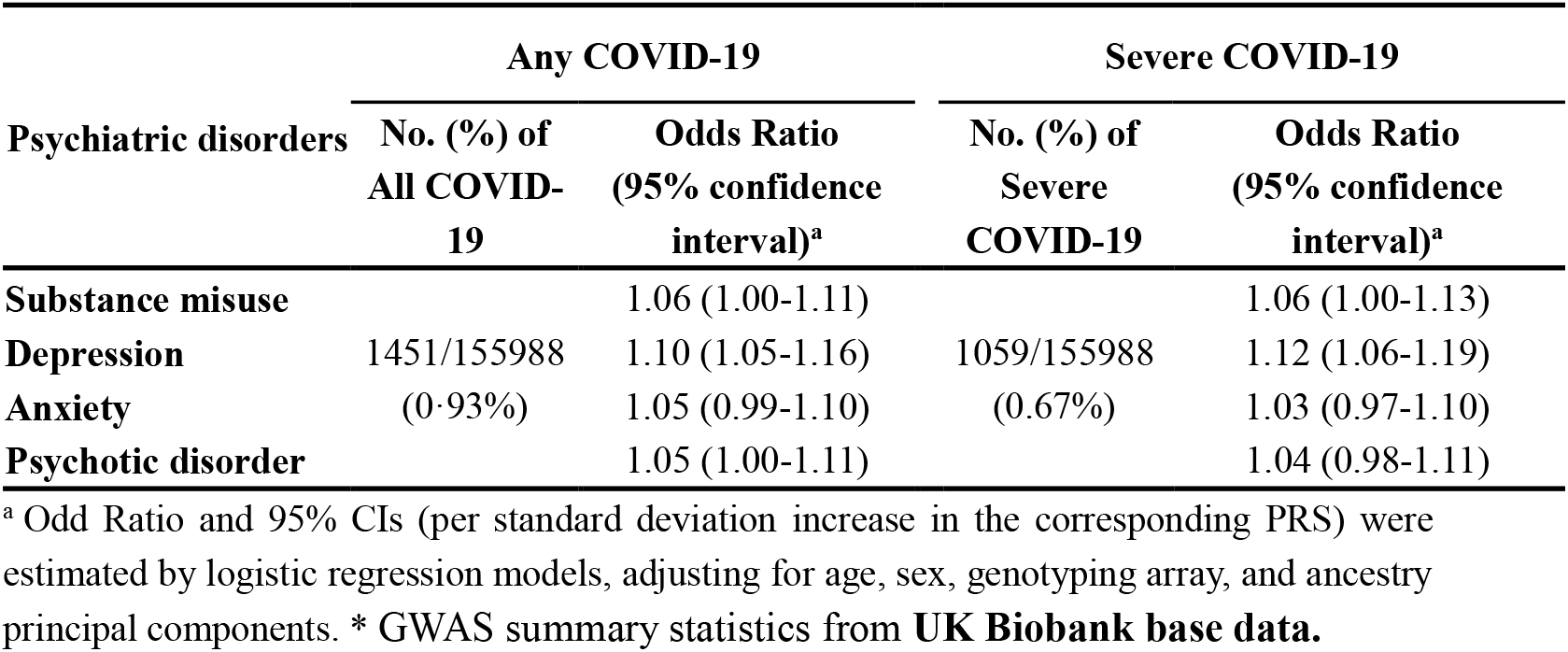
**The association between polygenic risk scores (PRSs) for psychiatric disorders and COVID-19\***

**Figure 3.**
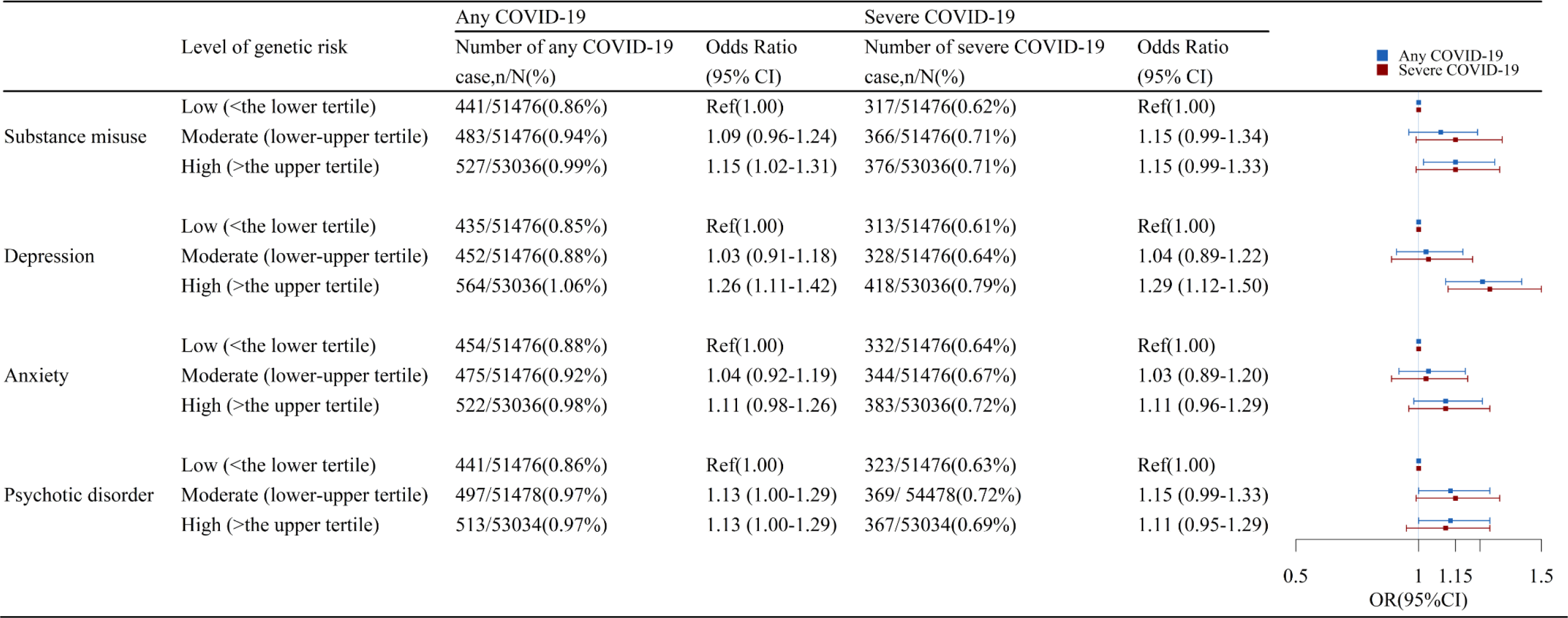
The association between categorized polygenic risk scores (PRSs) for psychiatric disorders and COVID-19 risk *. ^*^ Odd Ratio and 95% CIs were estimated by logistic regression models, adjusting for age, sex, genotyping array, and ancestry principal components GWAS summary statistics from UK Biobank base data.

The PRSs generated from publicly available GWAS summary statistics also yield significant correlations with its corresponding trait in UK Biobank data. Specifically, the results indicated better prediction accuracy for psychotic disorder but less accuracy for the other three studied psychiatric traits, compared to those generated based on GWAS of UK Biobank data (Supplementary Tables 2-5). The association analyses on PRS of each psychiatric disorder and COVID-19 revealed similar risk patterns as the main analyses, with somewhat attenuated estimates (Supplementary Table 7 and Supplementary Figure 3-4).

## Discussion

Based on the large-scale prospective cohort of UK Biobank with comprehensive phenotypic and genetic information, we confirmed that genetic predisposition to various psychiatric disorders, including depression, substance misuse and psychotic disorder, was associated to risk of COVID-19, especially severe COVID-19. To our knowledge, this is the first study to date that examined the relationship of genetic basis of psychiatric disorders with COVID-19; and our findings aid the interpretation of the reported phenotypic association of pre-pandemic psychiatric disorders with the risk of COVID-19 by demonstrating a role of genetic underlying mechanism in this link. The genetically driven susceptibility to COVID-19, especially to severe and fatal COVID-19, among individuals prone to psychiatric disorders underscores the need of heightened clinical awareness and medical care for this vulnerable population during COVID-19 pandemic. Further studies are however still warranted to verify these findings and to explore the functional mechanisms.

The hypothesis that a pre-existing psychiatric disorder may influence the susceptibility to infectious diseases has since long been discussed. Many previous studies have consistently reported an elevated risk of infection, including respiratory virus infections^25^, pneumonia^26^, sepsis^27^, and other life-threatening infections^3^, subsequent to the occurrence of psychiatric illness, particularly stress reaction and related psychiatric disorders. The proposed mechanisms for the observed associations include dysregulations in immune responses and inflammatory profile^28^, physiological alternations commonly accompany psychiatric presentations. Also, the possible changes in lifestyle, such as smoking and alcohol abuse^11^, following the occurrence of psychiatric disorders may have also contributed to the altered susceptibility to infections. More recently, a Danish national study suggested a genetic component in the association between psychiatric disorders and infections, by demonstrating a strong genetic link between having at least one psychiatric diagnosis and the occurrence of infection^12^. Furthermore, a GWAS analysis found that schizophrenia-associated genetic basics have important roles in immunity, providing support for the speculated genetic link between the immune system and schizophrenia^22^.

Since the outbreak of COVID-19, accumulating evidence, including our previous work^9^, has revealed that psychiatric disorders are associated with an elevated risk of COVID-19 and its related hospitalization and death^2,5–8^. Further, an excess risk of hospitalization for other infections was also observed at a comparable level to that of COVID-19, among individuals with pre-existing psychiatric disorders during the COVID-19 outbreak^9^. Therefore, it is reasonable to speculate that the increased susceptibility to COVID-19 and other infections may have shared etiologies, such as immune dysfunction^9^. Indeed, increased cytokine levels have been reported to be correlated with disease deterioration and fatal COVID-19^29^, implying a role of altered immune responses in the disease progression. Our study provides supportive data from the perspective of genetics, by indicating a genetic association between psychiatric disorder and COVID-19. A large proportion of the general population has a history of psychiatric disorders (17%-29%)^30^. The prevalence is even higher in the elderly population (46%)^31^ who are also more vulnerable to COVID-19^32^. The present findings highlight therefore the necessity of improved surveillance and the need of further explorations of possible interventions to reduce the risk of COVID-19 occurrence and disease deterioration among this vulnerable group.

Intriguingly, contrast to similar phenotypic associations of depression (OR = 1.62) and anxiety (OR = 1.60) with COVID-19^9^, the present genetic analysis revealed a differential importance of genetic component on these associations. Compared to depression, the increased susceptibility to COVID-19 among patients with anxiety seems to be less attributable to the shared genetic liability, and thereby may be more behavior- or environment-related. Different behavior and mobility patterns have indeed been reported among individuals with depression and anxiety, during COVID-19 pandemic^33,34^. Individuals with depression or negative emotion in response to COVID-19 outbreak tend to implement self-limiting mobility mode (i.e., restricting their daily mobility range)^33,35^, whilst individuals with anxiety were demonstrated to be less resilient to preventive measures (e.g., wearing a face mask, hand washing, social distancing, and self-isolation or quarantine) during the COVID-19 outbreak^36,37^.

The merits of the present study include the application of PRS for identifying genetic association between psychiatric disorder and COVID-19 in a large population, which corroborates the previously observed phenotypic association and sheds light on the underlying mechanisms. In addition, as population heterogeneity may undermine the prediction accuracy of PRS, not only due to the different ancestries but also differences in characteristics of populations such as differences in sex, age or socioeconomic status^18^, we used an internal base dataset to calculate the PRS of each psychiatric disorder, and then used such PRS to test the genetic association using a target dataset. Similar methods have been applied to investigate the association of genetic predisposition to asthma with COVID-19^13^. Furthermore, given that the similar analyses based on publicly available GWAS summary statistics corroborated the results of the main analyses, our findings seem robust to the choice of the GWAS summary statistics. Our study has several limitations. First, the genetic profile of stress-related disorders was not established due to limited sample size. We could therefore not examine the genetic association between stress-related disorders and COVID-19. Second, in selecting SNPs for PRS construction, relatively loose p-value thresholds were applied. For example, as determined by the presence of highest R square, the threshold was 0.5, 0.1 and 0.2 for substance misuse, depression, and anxiety, respectively, possibly resulting in noise from redundant loci and subsequently unclear impact on the studied association. Third, the UK Biobank participants are not representative of the general population in the UK^38^ and our genetic analysis is further limited to participants with white European ancestry which reduces the generalization of our findings to the whole UK population and other populations. Last, given that the PRS contains information from relatively common variants only, the impact of rare and low frequency genetic variants needs further investigation.

In conclusion, in the UK Biobank population, we found genetic predisposition to psychiatric disorders to be associated with increased risk of COVID-19, which may partially explain the observed phenotypic association between psychiatric disorders and COVID-19. Notably, the gene-driven susceptibility to COVID-19 among individuals prone to psychiatric disorders underscores the need of extra awareness and medical care for this vulnerable population. Further studies are needed to identify particular genetic variants with the aim to understand the underlying mechanisms, optimize risk stratification, and provide molecular targets for disease prevention.

## Data Availability

Data from the UK Biobank (http://www.ukbiobank.ac.uk/) are available to all researchers upon making an application. Part of this research was conducted using the UK Biobank Resource under Application 54803

## Acknowledgments

We would like to thank the team members and colleagues involved in West China Biomedical Big Data Center-UK Biobank project for their support.

## Contributions

HS, CS, UAV and FF were responsible for the study concept and design. YH, ZY, YS, and YQ did the data and project management. WC, YZ, HY, YC, CH did the data cleaning and analysis. WC, YZ, HY, CS, FF, UAV, DL and HS interpreted the data. WC, YZ, CS, UAV, and HS drafted the manuscript. All the authors approved the final manuscript as submitted and agree to be accountable for all aspects of the work.

## Declaration of interests

We declare no competing interests.

## Funding

This work is supported by the National Science Foundation of China (No. 81971262 to HS), West China Hospital COVID-19 Epidemic Science and Technology Project (No. HX-2019-nCoV-014 to HS), and Sichuan University Emergency Grant (No. 2020scunCoVyingji10002 to HS), and NordForsk grant (105668 to UV and FF).

## Ethical approval

The UK Biobank has full ethical approval from the NHS National Research Ethics Service (reference number: 16/NW/0274), and this study was also approved by the biomedical research ethics committee of West China Hospital (reference number: 2020.661).

## Notes

### Competing Interest Statement

The authors have declared no competing interest.

### Author Declarations

This present study was approved by the biomedical research ethics committee of West China Hospital (2020.661)

