## Supplemental Tables and Figures for "Genetic predisposition to psychiatric disorders and risk of COVID-19"

**Supplementary Table**

| **Diagnose** | **ICD-10** | **ICD-9** |
| --- | --- | --- |
| Substance misuse | F10-F19 | 291, 303-305 |
| Depression | F32-F33 | 2961, 3004, 311 |
| Anxiety | F40-F41 | 3000, 3002 |
| Psychotic disorder | F20-F29 | 295, 297, 298 |
| Stress-related disorder | F43 | 308, 309 |
| COVID-19 | U07.1, U07.2 | - |

**Supplementary Table 1 International Classification of Disease (ICD) codes, ninth (ICD-9) and tenth (ICD-10) revisions for diagnoses used in this study**

**Supplementary Table 2 Associations between substance misuse PRSs and the risk of substance misuse at different p value thresholds**

| Pt | **GWAS summary statistics from UK Biobank base data** | | | |  | **GWAS summary statistics from ICC** | | | |
| --- | --- | --- | --- | --- | --- | --- | --- | --- | --- |
|  | N_SNP | OR (95% CI) | R^2^ | p value |  | N_SNP | OR (95% CI) | R^2^ | p value |
| Pt5e-08 | - | - | - | - |  | - | - | - | - |
| Pt1e-06 | 1 | 1.00 (0.98-1.02) | 2.154% | 0.881 |  | 2 | 1.01 (0.99-1.02) | 1.574% | 0.507 |
| Pt1e-04 | 35 | 1.04 (1.01-1.06) | 2.170% | 1.36×10^-3^ |  | 79 | 1.02 (1.00-1.03) | 1.577% | 0.049 |
| Pt0.001 | 310 | 1.07 (1.05-1.10) | 2.211% | 7.03×10-^10^ |  | 639 | 1.01 (1.00-1.03) | 1.576% | 0.121 |
| Pt0.05 | 6861 | 1.16 (1.13-1.19) | 2.418% | 8.17×10^-40^ |  | 16859 | 1.05 (1.03-1.06) | 1.597% | 8.87×10^-8^ |
| Pt0.1 | 11379 | 1.17 (1.14-1.20) | 2.449% | 2.6×10^-44^ |  | 28634 | 1.05 (1.03-1.07) | 1.603% | 2.23×10^-9^ |
| Pt0.2 | 18267 | 1.17 (1.14-1.19) | 2.442% | 2.08×10^-43^ |  | 47172 | 1.05 (1.03-1.07) | 1.600% | 1.38×100^-8^ |
| Pt0.3 | 23649 | 1.17 (1.14-1.19) | 2.436% | 1.86×10-^42^ |  | 62092 | 1.05 (1.03-1.06) | 1.599% | 3.54×10^-8^ |
| Pt0.4 | 27989 | 1.17 (1.14-1.19) | 2.441% | 2.65×10^-43^ |  | 74199 | 1.05 (1.03-1.07) | 1.602% | 4.06×10^-9^ |
| Pt0.5 | 31543 | 1.17 (1.14-1.19) | 2.436% | 1.87×10^-42^ |  | 84034 | 1.05 (1.03-1.07) | 1.601% | 7.09×10^-9^ |

OR were adjusting for age, sex, genotyping array, and ancestry principal components. Pt was p value threshold. N_SNP was the number of SNP used predictive model. R^2^ was Nagelkerke’s squared (R square). PRS = polygenic risk score. GWAS = Genome wide association study. ICC = International Cannabis Consortium. – No results in this fields.

**Supplementary Table 3 Associations between depression PRSs and the risk of depression at different** **p value thresholds**

| Pt | **GWAS summary statistics from UK Biobank base data** | | | |  | **GWAS summary statistics from PGC** | | | |
| --- | --- | --- | --- | --- | --- | --- | --- | --- | --- |
|  | N_SNP | OR (95% CI) | R^2^ | p value |  | N_SNP | OR (95% CI) | R^2^ | p value |
| Pt5e-08 | - | - | - | - |  | 1 | 1.04 (1.02-1.05) | 0.984% | 2.62×10^-5^ |
| Pt1e-06 | 1 | 1.00 (0.98-1.02) | 1.048% | 0.923 |  | 9 | 1.04 (1.02-1.05) | 0.985% | 1.53×10^-5^ |
| Pt1e-04 | 51 | 1.02 (1.00-1.05) | 1.054% | 0.047 |  | 239 | 1.07 (1.05-1.08) | 1.019% | 1.38×10^-14^ |
| Pt0.001 | 310 | 1.05 (1.03-1.07) | 1.075% | 2.63×10^-5^ |  | 1225 | 1.11 (1.10-1.13) | 1.110% | 2.54×10^-38^ |
| Pt0.05 | 6910 | 1.11 (1.08-1.13) | 1.172% | 2.84×10^-19^ |  | 23552 | 1.17 (1.15-1.19) | 1.271% | 3.65×10^-80^ |
| Pt0.1 | 11415 | 1.12 (1.09-1.14) | 1.193% | 2.57×10^-22^ |  | 39286 | 1.18 (1.16-1.20) | 1.292% | 1.33×10^-85^ |
| Pt0.2 | 18359 | 1.13 (1.11-1.16) | 1.230% | 1.29×10^-27^ |  | 65088 | 1.18 (1.16-1.20) | 1.304% | 1.61×10^-88^ |
| Pt0.3 | 23784 | 1.14 (1.12-1.17) | 1.258% | 1.65×10^-31^ |  | 86406 | 1.18 (1.16-1.20) | 1.301% | 8×10^-88^ |
| Pt0.4 | 28167 | 1.15 (1.12-1.17) | 1.270% | 2.61×10^-33^ |  | 104418 | 1.18 (1.16-1.20) | 1.303% | 2.1×10^-88^ |
| Pt0.5 | 31716 | 1.14 (1.12-1.17) | 1.262% | 3.51×10^-32^ |  | 119795 | 1.18 (1.16-1.20) | 1.304% | 1.55×10^-88^ |

OR were adjusting for age, sex, genotyping array, and ancestry principal components. Pt was p value thresholds. N_SNP was the number of SNP in the predictive model. R^2^ was Nagelkerke’s squared (R square). PRS = polygenic risk score. GWAS = Genome wide association study. PGC = Psychiatric Genomics Consortium. – No results in this fields.

**Supplementary Table 4 Associations between anxiety PRSs and the risk of anxiety at different p value thresholds**

| Pt | **GWAS summary statistics from UK Biobank base data** | | | |  | **GWAS summary statistics from ICC** | | | |
| --- | --- | --- | --- | --- | --- | --- | --- | --- | --- |
|  | N_SNP | OR (95% CI) | R^2^ | p value |  | N_SNP | OR (95% CI) | R^2^ | p value |
| Pt5e-08 | 1 | 1.02 (0.99-1.04) | 1.290% | 0.263 |  | 1 | 0.99 (0.97-1.01) | 1.271% | 0.292 |
| Pt1e-06 | 1 | 1.03 (1.00-1.06) | 1.297% | 0.025 |  | 1 | 0.99 (0.97-1.01) | 1.271% | 0.292 |
| Pt1e-04 | 37 | 1.01 (0.99-1.04) | 1.289% | 0.359 |  | 86 | 1.01 (0.99-1.03) | 1.271% | 0.32 |
| Pt0.001 | 266 | 1.06 (1.03-1.09) | 1.325% | 9.82×10^-6^ |  | 621 | 1.03 (1.01-1.05) | 1.281% | 1.13×10^-3^ |
| Pt0.05 | 6630 | 1.10 (1.08-1.13) | 1.399% | 2.61×10^-14^ |  | 16960 | 1.05 (1.04-1.07) | 1.302% | 2.95×10^-8^ |
| Pt0.1 | 11087 | 1.11 (1.09-1.14) | 1.419% | 1.45×10^-16^ |  | 28876 | 1.05 (1.03-1.07) | 1.302% | 4.07×10^-8^ |
| Pt0.2 | 18067 | 1.12 (1.09-1.14) | 1.422% | 5.71×10^-17^ |  | 47277 | 1.05 (1.03-1.07) | 1.297% | 3.26×10^-7^ |
| Pt0.3 | 23472 | 1.12 (1.09-1.15) | 1.425% | 2.88×10^-17^ |  | 61605 | 1.05 (1.03-1.07) | 1.301% | 5.74×10^-8^ |
| Pt0.4 | 27943 | 1.12 (1.09-1.15) | 1.436% | 1.75×10^-18^ |  | 72743 | 1.05 (1.03-1.07) | 1.301% | 4.79×10^-8^ |
| Pt0.5 | 31497 | 1.12 (1.09-1.15) | 1.440% | 6.15×10^-19^ |  | 82028 | 1.05 (1.03-1.07) | 1.301% | 6.35×10^-8^ |

OR were adjusting for age, sex, genotyping array, and ancestry principal components. Pt was p value cutoff thresholds. N_SNP was the number of SNP in the predictive model. R^2^ was Nagelkerke’s squared (R square). PRS = polygenic risk score. GWAS = Genome wide association study. PGC = Psychiatric Genomics Consortium.

**Supplementary Table 5 Associations between psychotic disorder PRSs and the risk of psychotic disorder at different p value thresholds**

| Pt | **GWAS summary statistics from UK Biobank base data** | | | |  | **GWAS summary statistics from ICC** | | | |
| --- | --- | --- | --- | --- | --- | --- | --- | --- | --- |
|  | N_SNP | OR (95% CI) | R^2^ | p value |  | N_SNP | OR (95% CI) | R^2^ | p value |
| Pt5e-08 | - | - | - | - |  | 112 | 1.12 (1.04-1.20) | 1.782% | 3.07×10^-3^ |
| Pt1e-06 | 1 | 1.07 (0.98-1.17) | 2.804% | 0.154 |  | 203 | 1.17 (1.08-1.26) | 1.857% | 4.84×10^-5^ |
| Pt1e-04 | 32 | 0.98 (0.89-1.07) | 2.775% | 0.658 |  | 1059 | 1.34 (1.24-1.44) | 2.233% | 8.14×10^-14^ |
| Pt0.001 | 229 | 0.97 (0.88-1.06) | 2.780% | 0.476 |  | 3127 | 1.42 (1.32-1.53) | 2.523% | 6.58×10^-21^ |
| Pt0.05 | 6263 | 1.07 (0.98-1.17) | 2.810% | 0.128 |  | 31195 | 1.66 (1.54-1.78) | 3.467% | 2.63×10^-44^ |
| Pt0.1 | 10729 | 1.13 (1.03-1.23) | 2.891% | 6.04×10^-3^ |  | 48505 | 1.65 (1.54-1.77) | 3.442% | 8.4×10^-44^ |
| Pt0.2 | 17811 | 1.12 (1.03-1.22) | 2.883% | 7.39×10^-3^ |  | 75739 | 1.66 (1.55-1.79) | 3.480% | 9.24×10^-45^ |
| Pt0.3 | 23332 | 1.13 (1.04-1.23) | 2.894% | 4.84×10^-3^ |  | 97661 | 1.68 (1.57-1.80) | 3.551% | 1.6×10^-46^ |
| Pt0.4 | 27779 | 1.14 (1.05-1.24) | 2.912% | 2.31×10^-3^ |  | 116236 | 1.68 (1.57-1.81) | 3.562% | 8.75×10^-47^ |
| Pt0.5 | 31395 | 1.15 (1.06-1.25) | 2.927% | 1.23×10^-3^ |  | 131898 | 1.67 (1.56-1.80) | 3.521% | 7.98×10^-46^ |

OR were adjusting for age, sex, genotyping array, and ancestry principal components. Pt was p value thresholds. N_SNP was the number of SNP in the predictive model. R^2^ was Nagelkerke’s squared (R square). PRS = polygenic risk score. GWAS = Genome wide association study. PGC = Psychiatric Genomics Consortium. – No results in this fields.

**Supplementary Table 6 Associations between stress-related disorder PRSs and the risk of stress-related disorder at different p value thresholds**

| Pt | **GWAS summary statistics from UK Biobank base data** | | | |
| --- | --- | --- | --- | --- |
|  | N_SNP | OR (95% CI) | R^2^ | p value |
| Pt5e-08 | - | - | - | - |
| Pt1e-06 | 18 | 0.97 (0.87-1.09) | 4.718% | 0.596 |
| Pt1e-04 | 462 | 0.88 (0.78-0.98) | 4.823% | 0.025 |
| Pt0.001 | 2432 | 0.90 (0.80-1.00) | 4.792% | 0.055 |
| Pt0.05 | 64236 | 0.95 (0.85-1.06) | 4.733% | 0.324 |
| Pt0.1 | 120096 | 0.95 (0.85-1.06) | 4.733% | 0.321 |
| Pt0.2 | 215678 | 0.96 (0.86-1.08) | 4.721% | 0.513 |
| Pt0.3 | 297378 | 0.97 (0.87-1.08) | 4.719% | 0.578 |
| Pt0.4 | 364888 | 0.97 (0.87-1.09) | 4.717% | 0.632 |
| Pt0.5 | 423862 | 0.98 (0.88-1.09) | 4.715% | 0.685 |

OR were adjusting for age, sex, genotyping array, and ancestry principal components. Pt was p value thresholds. N_SNP was the number of SNP in the predictive model. R^2^ was Nagelkerke’s squared (R square). PRS = polygenic risk score. GWAS = Genome wide association study.

**Supplementary Table 7 The association between polygenic risk scores (PRSs) for psychiatric disorders and COVID-19***

| **Psychiatric disorders** | **Any COVID-19** | |  | **Severe COVID-19** | |
| --- | --- | --- | --- | --- | --- |
|  | **No. (%) of Any COVID-19** | **Odds Ratio**  **(95% confidence interval)^a^** |  | **No. (%) of Severe COVID-19** | **Odds Ratio**  **(95% confidence interval)^a^** |
| Substance misuse | 1451/287240 (0.51%) | 1.05 (1.00-1.11) |  | 1059/287240 (0.37%) | 1.05 (0.99-1.12) |
| Depression |  | 1.04 (0.98-1.09) |  |  | 1.05 (0.99-1.11) |
| Anxiety |  | 1.04 (0.99-1.10) |  |  | 1.08 (1.02-1.15) |
| Psychotic disorder |  | 1.03 (0.98-1.09) |  |  | 1.05 (0.99-1.11) |

a Odd Ratio and 95% CIs (per standard deviation increase in the corresponding PRS) were estimated by logistic regression models, adjusting for age, sex, genotyping array, and ancestry principal components. *GWAS summary statistics from **publicly available GWAS summary statistics.**

Included participants

346,554

97,059,328

Restricted to the autosomal biallelic SNPs and imputation accuracy score > 0.1

SNPs with call rate > 98%, a minor allele frequency > 0.01, and Hardy–Weinberg equilibrium (p < 10^−6^)

Participants with genotyping rate > 98%

Participants with heterozygosity within ±3 SD from mean

**Participants and SNPs passed GWAS QC steps**

340,632

**340,632**

4,813,217

86,178,275

**4,813,217**

**SNPs**

**Participants**

346,554

**Supplementary Figure 1 Flowchart of the entire GWAS quality control process**

**A Substance misuse**

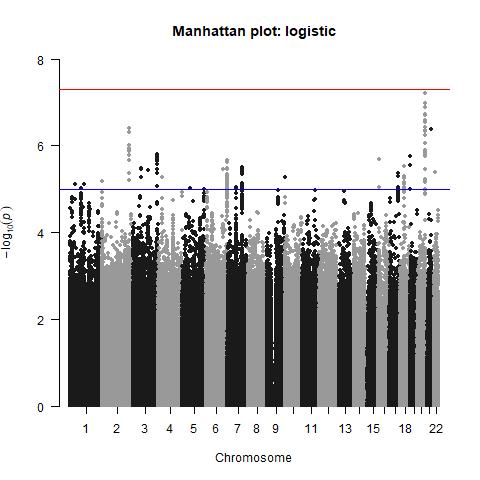

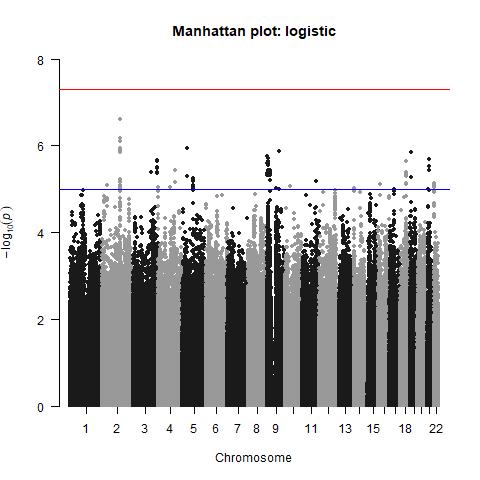

**B Depression**

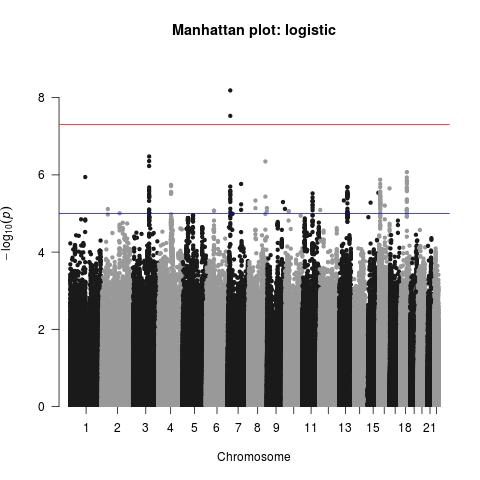

**C Anxiety**

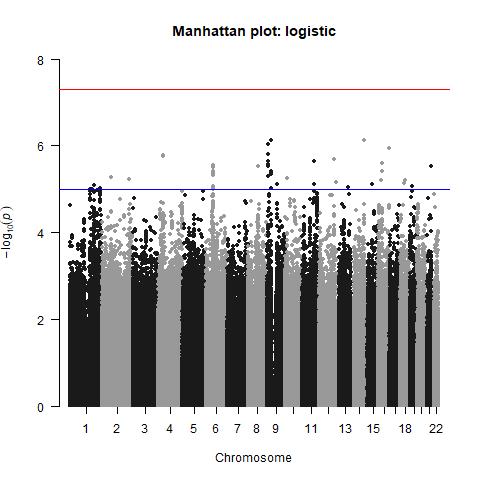

**D Psychotic disorder**

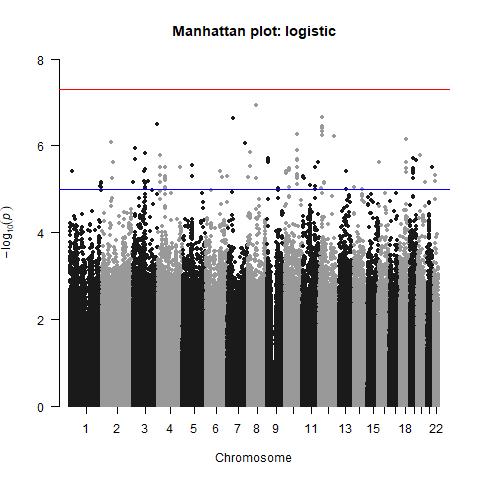

**E Stress-related disorder**

**Supplementary Figure 2 Manhattan of the SNP-Based GWAS of psychiatric disorders (**SNP=single-nucleotide polymorphism)

Variance explained by substance misuse PRS

Variance explained by depression PRS

Variance explained by anxiety PRS

Variance explained by psychotic disorder PRS

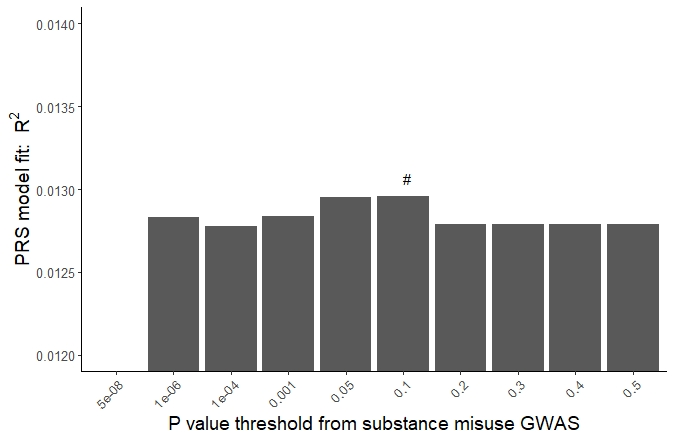

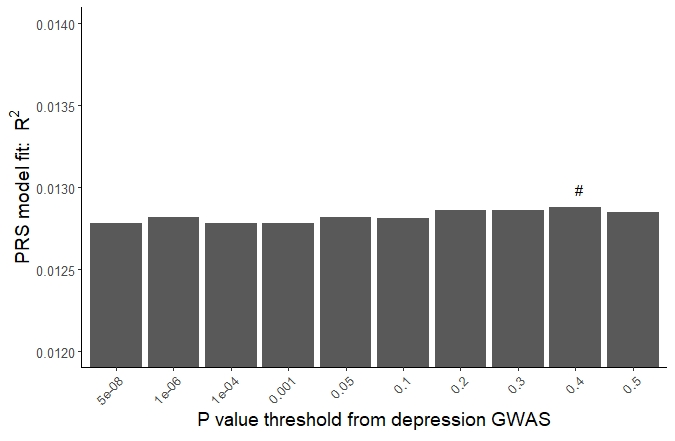

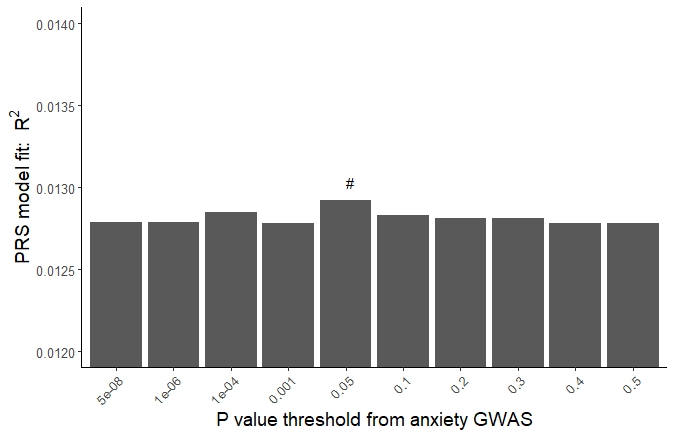

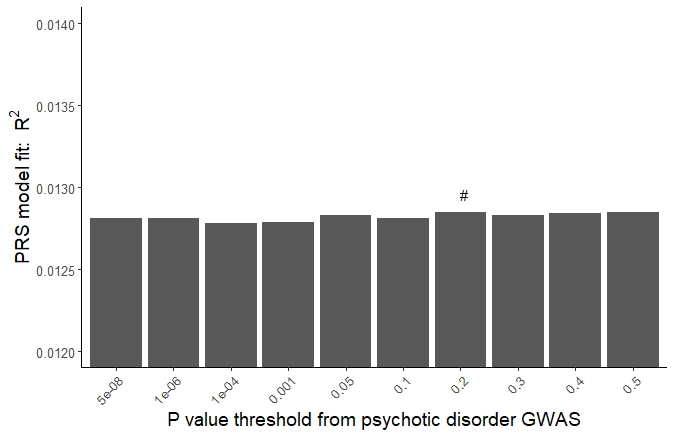

**Supplementary Figure 3 Variance explained for COVID-19 by polygenic risk scores (PRSs) for psychiatric disorder under 10 p value thresholds***

**#** The p value threshold with the highest variance explained was selected for subsequent analyses

*GWAS summary statistics from **publicly available GWAS summary statistics**

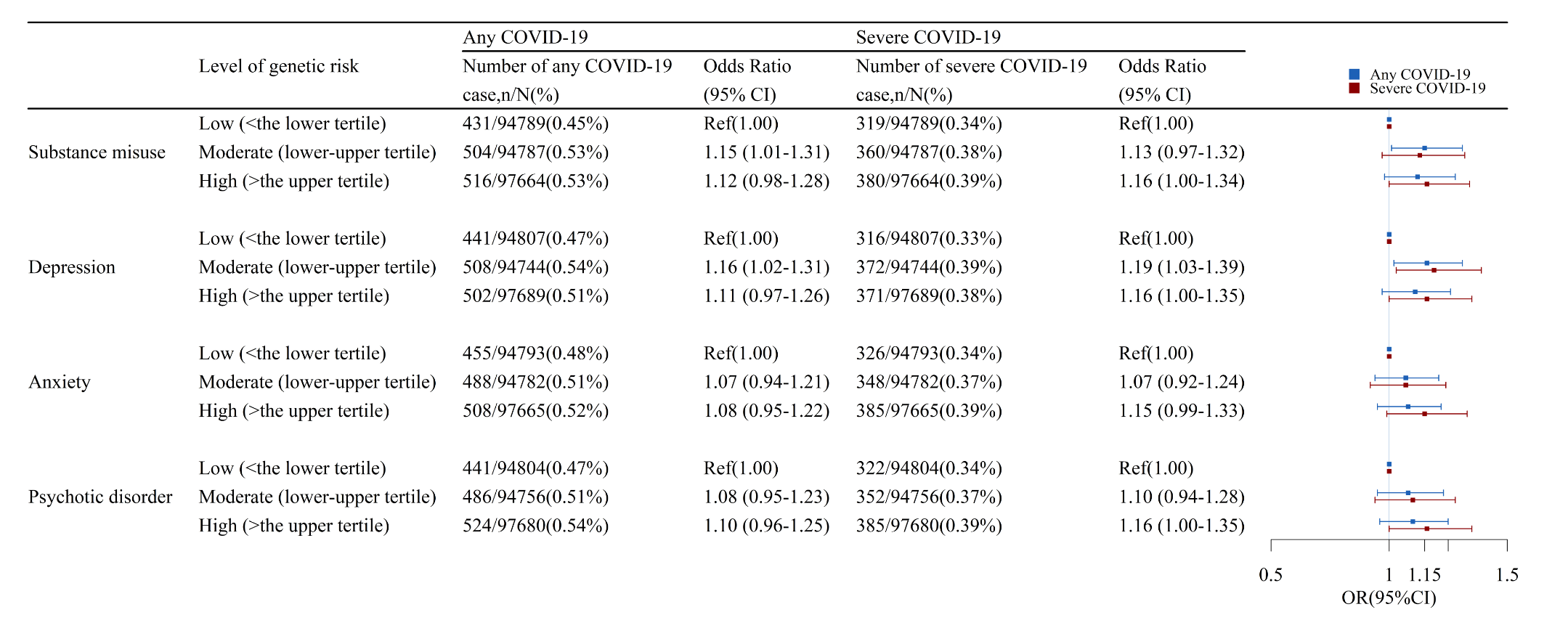

**Supplementary Figure 4 The association between categorized polygenic risk scores (PRSs) for psychiatric disorders and COVID-19 risk ***

* Odd Ratio and 95% CIs were estimated by logistic regression models, adjusting for age, sex, genotyping array, and ancestry principal components. GWAS summary statistics from **publicly available GWAS summary statistics**
